# NURSING PHILOSOPHY OF FAMILY SUPPORT FOR PATIENT CARE SKIZOFRENIA: A LITERATURE REVIEW

**DOI:** 10.1101/2024.12.20.24318753

**Authors:** Aristina Halawa, Ah. Yusuf, Moses Glorino Rumambo Pandin

**Affiliations:** Doctor of Nursing, Faculty of Nursing Universitas Airlangga, Jalan Dr. Ir.H. Soekarno, Mulyorejo, Kec. Mulyorejo, Surabaya, East Java 60115; Faculty of Nursing Universitas Airlangga, Jalan Dr. Ir. H. Soekarno, Mulyorejo, Kec. Mulyorejo, Surabaya, East Java 60115; Faculty of Humanities Universitas Airlangga, Jalan Dr. Ir. H. Soekarno, Mulyorejo, Kec. Mulyorejo, Surabaya, East Java 60115; William Booth College of Health Sciences, Jalan Cimanuk no 20, Wonokromo Surabaya, East Java 60241

**Keywords:** Family support, care, schizophrenia

## Abstract

**Introduction:** The emergence of deinstitutionalisation of people with mental disorders has led to the burden of patient care being shifted to families. Patients with schizophrenic disorders who live with their families account for up to 50%. Families play an important role in the lives of individuals living with mental illnesses such as schizophrenia. Caregivers often face economic burdens and challenges in managing daily household tasks, which can exacerbate stress and reduce their overall well-being. This family support is lacking as family caregivers often experience significant emotional and physical stress due to the high demands of caregiving. If an individual receives emotional, informational, or instrumental support from their family then they will likely have greater confidence, hope, and optimism to pursue recovery and live a fulfilling and meaningful life.

**Methods:** This research uses a literature review using the PICOS framework. The literature for the originality of this research uses articles in English from 4 databases: Science direct, Scopus, Sage and EBSCOhost. The keywords used in the literature search were: “family support” AND family AND support AND schizophrenia. The search was limited to publications from 2019-2024 and the articles retrieved were full articles not review articles. The strategy used to search for articles used the PICOS framework. Then selected using PRISMA diagram and obtained ten articles.

**Results:** There are 10 articles obtained and all of them explain about family support in caring for schizophrenia patients both from factors that affect family support, forms of family support and the impact of family support on caregivers or family members and patients. Thus, family support is very important for the welfare of caregivers and for patients who experience schizophrenia disorders.

**Conclusion:** Family support is essential in the care of patients with schizophrenia. There are several barriers that prevent families from accessing necessary resources and support, including stigma, lack of information, and limited contact with the healthcare team. Family Support provided to caregivers as well as to patients provides better outcomes for patients including increased independence and improved daily functioning, reduced caregiver burden and improved caregiver ability to provide care.

## Introduction

Schizophrenia is a chronic mental disorder that can pose a major threat to individuals, families and society. The rise of deinstitutionalisation of people with mental illness shifts the burden of care to families.( Mbadugha et al., 2023) Families play an important role in the lives of individuals living with mental illnesses such as schizophrenia. Schizophrenia is a highly disabling mental illness and affects the functioning of family caregivers (Fatima & Tariq, 2022). The presence of psychotic symptoms in adults with schizophrenia requires increased control and family support to prevent the risk of aggressive behaviour (Lekganyane, 2020) Patients with schizophrenia disorder who live with their families reach 50%. The responses of families who have a family member with schizophrenia include: burden of care, fear and shame of signs and symptoms of illness, uncertainty about the course of illness, lack of social support, and stigma. The stigma of mental illness is a social phenomenon that occurs in society about people who cannot accept individuals who experience mental illness because they have unnatural behaviour or character, have a deviant social identity, so this makes people tend to discriminate (Shamsaei et al., 2015) According to the family psychology recovery model one of the factors that influence recovery from mental illness is family support.(Mabunda, 2024) Family support is defined as the attitudes and behaviours of the family in supporting its members (House et al, 1985), which can take the form of emotional support (e.g., care and comfort), informational support (e.g., guidance and advice), or instrumental support (e.g., practical and tangible assistance (Chan et al., 2023). If an individual receives emotional, informational, or instrumental support from their family then they will likely have greater confidence, hope, and optimism to pursue recovery and live a fulfilling and meaningful life. (Sánchez et al., 2019)

There are several barriers that prevent families from accessing necessary resources and support, including stigma, lack of information, and limited contact with the health team. (Drapalski et al., 2009). This family support is lacking as family caregivers often experience significant emotional and physical stress due to the high demands of caregiving. This stress can lead to reduced quality of life and mental health issues for the carers themselves.(Chen et al., 2019). Caregivers often face economic burdens and challenges in managing daily household tasks, which can exacerbate stress and reduce their overall well-being.(Sustrami, Yusuf, Fitryasari, Efendi, et al., 2023)

Schizophrenia sufferers worldwide are estimated to be 29 million people (Chan, 2011). Higher rates of premature mortality were reported for patients with schizophrenia, and it was found that up to 28% of these experienced early death by suicide. (Galletly et al, 2016). Basic Health Research in 2018 stated that the prevalence of schizophrenia in Indonesia reached 6.7 per 1,000 population which increased from 2013 of 1.7 per 1,000 population. The prevalence of people with mental disorders in East Java Province in severe mental disorders (psychosis / schizophrenia) has data of 6.4 ‰ and the prevalence of people with mild mental disorders or patients with mental emotional disorders with symptoms, such as: depression and anxiety of 4% for ages 15 years and over or around 14 million population.(Sustrami, Yusuf, Fitryasari, Suhardiningsih, et al., 2023). Based on data from Menur Surabaya Mental Hospital, it was found that the number of schizophrenia patients has increased from year to year. This can be seen from the data found, namely in 2022 there were 20,890 while in 2023 there were 21,028 schizophrenia patients.

Improvement in schizophrenia is strongly linked to family involvement in the schizophrenic’s life. Family members can ease the difficulties of this serious mental illness in ways that people outside the family system cannot(He et al., 2021). Families are very important in caring for schizophrenia patients in the community, as many patients cannot be hospitalised due to limited numbers and availability of facilities, so schizophrenia patients are cared for in the family as the smallest unit of society. Data on patients with severe mental disorders in Indonesia are mostly in the community rather than in hospitals(Iswanti et al., 2023a) Families living with schizophrenia patients will experience heavy demands because they take up a lot of time to care for and provide support to make the patient feel better. The stigma associated with schizophrenia has a negative effect on family life, leading to a desire to physically and socially withdraw and limit socialising with the community. Negative treatment given to family members who care for mentally ill patients in the form of rejection, denial and exclusion.(Varghese et al., 2017) The high stigma experienced by families can hinder the treatment process and access to health services, which ultimately worsens the patient’s health condition.(He et al., 2021) The impact experienced by the family due to a family member with mental illness includes the family experiencing a sense of discomfort, frustration, anxiety, despair, grief, fatigue and helplessness due to the loss of their routine time during the care and treatment of patients, resulting in family rejection of patients who tend to blame people with mental disorders so that the lack of support for patients.(Brandt et al., 2022) Lack of family support can increase the emotional and psychological burden on patients and carers. Family carers often experience significant stress, financial burden, and social isolation when they do not receive adequate support.(Azman et al., 2019) Inadequate family involvement can negatively impact the quality of mental health care provided. Nurses have reported that poor family contact affects the provision of quality care, indicating that family support is essential for effective mental health management.(Mabunda, 2024).

Family support can be improved in several ways, with specialised training for health professionals on family engagement improving their ability to engage families effectively. Training can include communication skills, understanding family dynamics, and strategies to engage families in care planning.(Muddle et al., 2024). Engaging families in structured conversations with mental health professionals can assist in understanding and meeting their needs, promote healing, and reduce suffering.(Moen et al., 2021)

## Materials and Methods

This research uses a literature review using the PICOS framework. The literature for the originality of this research uses articles in English from 4 databases: Science direct, Scopus, Sage and EBSCOhost. The keywords used in the literature search were: “family support” AND family AND support AND schizophrenia. The search was limited to publications from 2019-2024 and the articles retrieved were full articles not review articles. Then selected using the PRISMA diagram and obtained ten articles.

**Figure 1.**
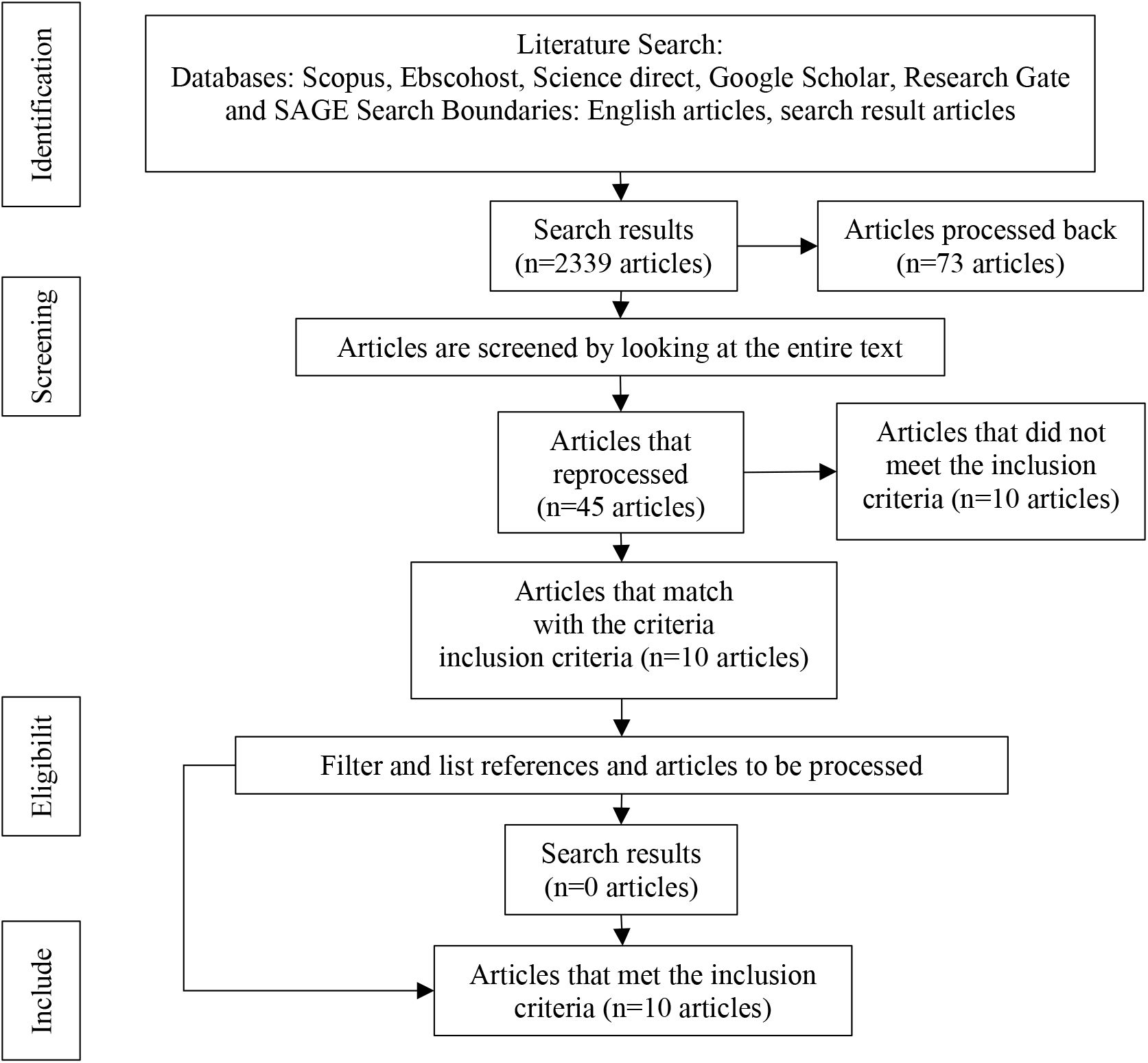
Prism Flow Diagram

**Table 1.** Journal Article Analysis Results.

| No | Research Title | Research Methods | Research Results |
| --- | --- | --- | --- |
| 1. | Family support and quality of life of schizophrenia patients. (Setiawati et al., 2021) | **Design:** Cross-sectional<br>**Sample:** Sample size 161 randomly selected outpatients in 13 community health centers<br>**Variables:** family support and quality of life and adherence.<br>**Instruments**<br>Friedman family support questionnaire and WHO quality of life questionnaire (WHOQOL-BREF).<br>Medication adherence was assessed using the Morisky Medication Adherence Scale (MMAS).<br>**Analysis:** logistic regression | Respondents had a mean age of 45 years, were mostly male, had completed high school, were mostly unemployed and unmarried. Instrumental support (AOR=3.177; 95%CI 1.01-9.91) and appraisal support (AOR=7.620; 95%CI 2.83-20.4) were significantly associated with QOL. In contrast, there was no significant association between emotional support (AOR=1.345; 95%CI 0.46-3.88) and informational support (AOR=2.515; 95%CI 0.85-7.42) on QOL. |
|  |  |  | Employment, being married and not experiencing relapse were significantly associated with QOL. Instrumental support and appraisal support are important factors in determining the quality of life of people with schizophrenia. |
| 2. | The burden, support and needs of primary family caregivers of people experiencing schizophrenia in Beijing communities: A qualitative study(Chen et al., 2019) | **Design:** Qualitative research design<br>**Sample:** 20 family caregivers of schizophrenia patients<br>**Variables:** explore caregivers' perspectives on caregiver burden, support and further needs (1) what difficulties do you encounter you faced when caring for the patient? (2) what support or resources have you obtained? (3) what further support do you need to provide better care for the patient? better care for the patient?"<br>**Instruments:** face-to-face and semi-structured in-depth interviews<br>**Analysis:** In the analysis process, Colaizzi's 7-step analysis phenomenological data method | Most of the participants reported that they suffered from heavy life burdens and had negative experiences in terms of getting social support, and they emphasis that they needed more support. Economic burdens and daily household chores, limited social communication, and psychological distress were the main burdens. Support including financial, medical and informational and educational support did not meet the needs of caregivers and patients. More financial support, respect, and rehabilitation institutions were reported. More were reported as needs of the carers. |
| 3. | Family caregivers' perspectives on barriers to caring for patients with schizophrenia: A descriptive qualitative study(Asgari et al., 2023) | **Design:** Qualitative research design<br>**Sample:** Sample size is 16 family caregivers of schizophrenia patients Sampling using purposive sampling.<br>**Variables:** Barriers to the care of patients with Schizophrenia<br>**Instruments:** Semi-structured in-depth interview<br>**Analysis:** This study uses a content analysis approach, which aims to explore the barriers experienced by family caregivers in providing care to patients with schizophrenia. | The findings are classified into three main categories.<br>1.The category "Inefficiency of Support Resources" includes the subcategories "Inadequate support by Family Members," "Inadequate Support by Healthcare System," and "Financial Hardship."<br>2.The category "Limited Public Knowledge of Mental Disorders" includes the subcategories "Social Stigmatization" and "Social Rejection."<br>3.The category "destructive nature of schizophrenia" includes the subcategories "Gradual Loss of Ability" and "Gradual Patient Passivity." |
| 4. | Family support for Persons with schizophrenia physical after restraint and | **Design:** : qualitative with a phenomenological approach<br>**Sample:** Sample size of 10 participants using purposive sampling technique | This study identified three themes, namely:<br>(1) supervision of drug use, (2) provision of sustainable and optimal care, and (3) |
|  | confinement (Purba et al., 2020) | **Variables:** family support needed by people with schizophrenia after experiencing restraint or confinement<br>**Instruments:** in-depth interviews with the researcher as an primary instrument<br>**Analysis:** Data was analyzed using seven stages in the Colaizzi method | empowerment of people with schizophrenia. Active participation of the family in the treatment and care of the patient is very important. Families are the main source of support for people with schizophrenia. They feel valued and dignified with attention and acceptance from the family. Social support from family can help them to adhere to taking medication, make patients a dignified person, empower patients and prevent relapse and overcome stigma. |
| 5. | Correlation Family Support on Independence of Patients Schizophrenic Activities Daily Living (ADL) (Rohmi et al., 2020) | **Design:** The design used was correlational with a cross-sectional approach.<br>**Sample:** Sample size of 70 families who have family members with schizophrenia.<br>**Variables:** The variables in this study are family support (instrumental, information, appreciation, appraisal) and ADL independence in people with schizophrenia<br>**Instruments:** family support and Independence Frequency Distribution (Index Katz) for people with Schizophrenia<br>**Analysis:** statistical test using Spearman Rank | Based on the results of statistical tests using Spearman Rank shows the $\rho$ value for instrumental family support (0.020), informational family support (0.031), appreciation family support (0.021), and appraisal family support (0.021) and valuation family support (0.021). Full family support is needed and given to patients, especially schizophrenia patients. Comprehensive family support can be provided if the caregiver is a nuclear family member who has good psychological closeness with the patient. |
| 6. | Knowledge, Perception, And Burden Of Family In Treating Patients With Schizophrenia Who Experience Relapse (Suryani et al., 2019) | **Design** This research uses a quantitative descriptive method<br>**Sample:** Sample size of 100 clients with schizophrenia with a sampling technique that is consecutive technique<br>**Variables:** Knowledge, family perception and family burden<br>**Instruments:** The knowledge and perceptions of the family questionnaire were developed by the researcher based on various relevant theoretical sources. The knowledge questionnaire measured the concept of schizophrenia and how to treat it. handling of schizophrenia patients, while the instrument to measure the level of family burden uses the Zarit Burden | All patients with schizophrenia in this study experienced relapse, and based on the research it appears that family knowledge is inadequate in dealing with schizophrenia patients, as well as the burden of most families were at moderate and high levels, while the perception of perception of schizophrenia is good. |
|  |  | Interview which is used to measure family burden.<br>**Analysis:** multilevel modelling to test session level prediction and linear regression analysis for treatment level prediction. |  |
| 7. | The Social Support Perceived by Patients with Schizophrenia at Ali-Kamal Center in Sulaimani City, Kurdistan of Iraq. (Abdulkarim, 2024) | **Design:** Quantitative descriptive research<br>**Sample:** Sample size of 100 patients with schizophrenia recruited from outpatient psychiatry clinics. Inclusion criteria were Patients aged 18 years and above of both sexes and have been diagnosed with schizophrenia previously by a psychiatrist for more than 2 years and are currently in remission, compliant with medication can communicate and visit the outpatient psychiatry centers with family, patients with neurocognitive disorders, and in the critical phase were excluded which based on the psychiatrist.<br>**Variables:** Social support, demographic characteristics<br>**Instruments:** This questionnaire consists of three sections. The first section consists of the patient's sociodemographic data, including age, gender, marital status, economic status, education level, area of residence, and occupation. The second section includes clinical characteristics including disease duration and age of onset. The third section was the multidimensional scale of perceived social support (MSPSS).<br>**Analysis:** Statistical Package for the Social Sciences (SPSS) version 22 software was used for data analysis. The predetermined level of significance was set at | Low social support is felt by schizophrenia patients. Gender $P=0.05$; economic status, $P=0.04$; and education level $P=0.01$ were statistically significantly associated with low social support among the sample. This study has several implications for clinical practice and policy. First, the significant associations between some socio demographic factors and perceived social support underscore the importance of comprehensive treatment approaches that address not only the clinical symptoms of schizophrenia but also personal factors affecting patients. Second, the high value placed on social support by patients suggests that interventions aimed at strengthening social networks could be beneficial in managing schizophrenia. |
| 8. | Navigating care: family information needs and responsibilities in the context of schizophrenia caregiving (Fitryasari et al., 2024a) | **Design:** descriptive qualitative<br>**Sample:** The sample size was 20 families of patients with schizophrenia<br>**Variables:** Participants were asked to answer two main questions: "What information do families need from mental health | 1. For the theme Provide support for patient care. The support provided by the family is mainly in two ways, namely bringing the patient to control mental health services regularly and monitoring the patient to |
|  |  | services when caring for patients with schizophrenia?" and "What are the responsibilities of families in caring for patients with schizophrenia at home?"<br>**Instruments:** Data was collected using in-depth interviews and field notes<br>**Analysis:** The data was analyzed and interpreted based on the five steps of the content analysis technique. | always take medication regularly.<br>2. For the theme of meeting daily needs. The family felt that they were the ones who should be responsible for fulfilling the patient's daily needs because the patient was sick and needed help. The patient's needs are categorized into three: basic daily needs such as eating, drinking, and snacks; self-care needs such as bathing and changing clothes, which the family must always remind them of; and personal needs such as clothing and daily pocket money.<br>3. For the theme of helping with social skills. For some families, patients with mental illness have difficulty socializing with others, so the family must help gradually so that the patient can interact well with family.<br>4. For the theme of providing leisure activities. The family said they wanted to provide activities for the patient in their free time. Every day the patient needs to be active to avoid relapse. Some families try to facilitate patients who like art activities by buying musical instruments or painting tools and taking art lessons. The family also involves the patient in doing routine household tasks such as cleaning the house, cooking, and washing clothes. |
| 9. | Family and peer resources in relation to psychological condition in patients with paranoid schizophrenia (Izydorczyk et al., 2019) | **Design:** quasy experiment by using a control group.<br>**Sample:** The study group consisted of 201 individuals, 95 females and 106 males<br>**Variables:**<br>1) life experiences<br>2) difficult situations<br>3) positive beliefs, about trust and loyalty in the family<br>4) family and social resources | The results of the presented study confirmed a significant relationship between family resources and support from friends and factors describing the psychological state of patients with paranoid schizophrenia. Higher beliefs regarding mutual trust and respect among family members, pride and loyalty |
|  |  | 5) Sociodemographic variables are also controlled: age, gender, education, source of income, family type, family economic situation, parental education and living conditions.<br>6) Psychological conditions<br>**Instruments:** Clinical interviews, surveys created for the study, to measure life experiences and ways of coping with adverse life situations and their subjective outcomes for social functioning, The Family Strengths Scale, The Perceived Social Support Scale, The Berne Questionnaire of Subjective Well-Being, The Positive and Negative Syndrome Scale, The Mood Scale, and The General Health Questionnaire.<br>**Analysis:** Descriptions used numerical frequencies and percentages, means, and standard deviations. Kolmogorov-Smirnov test | towards the family, and competence and efficiency in managing life problems were associated with higher psychiatric symptomatology and higher self-rated mental health in the subjects. The results of the presented study identified a significant relationship between family resources (the sick individual's beliefs regarding family trust and loyalty and perceived family support) and the psychological state and dynamics of the course of schizophrenia. The study showed that indicators of family and social support also mediate the intensity of healing indicators and the decrease in illness indicators, and sources of support are critical to the course of illness. |
| 10 | Factors related to family's ability to care for schizophrenic patients(Iswanti et al., 2023a) | **Design:** correlational research.<br>**Sample:** The sample of this study was 135 families of schizophrenia in the outpatient polyclinic.<br>**Variables:** Resources, Emotional expression, Social support, Infrastructure, Health workers, family's ability to care for schizophrenia patients.<br>**Instruments:** The instruments in this study were questionnaires developed and modified from the experience caregiving instrument (ECI), the inner resources scale (SAS-I), the mental health inventory (MHI), the Berkeley Expressivity Questionnaire, the Barthel Index, the Caregiving Tasks in Caring for Adults with Mental Illness Scale (CTiCAMIS).<br>**Analysis:** Data was analyzed using multiple regression tests | The ability of families to care for schizophrenia patients can be predicted by resource factors, social environment. Family support, family emotional expression, infrastructure, and health workers. The strongest predictor factor is the resources owned by the family, which is 57.5%.<br>The results also show that there is a relationship between family emotional expression, infrastructure and health workers with the family's ability to care for schizophrenia patients, although these three variables are not predictors. The facilities and infrastructure available at the hospital as well as health workers who support families to provide good care for schizophrenia patients will be a source for families to improve their ability to care<br>for patients by accessing local health services. |

## Results and Discussion

Based on the results of the analysis of 10 journal articles describing family support in caring for schizophrenia patients, it was found in the study (Setiawati et al., 2021) (Chen et al., 2019) (Asgari et al., 2023) (Purba et al., 2020) (Rohmi et al., 2020) (Suryani et al., 2019) (Abdulkarim, 2024) (Fitryasari et al., 2024a) (Izydorczyk et al., 2019) (Iswanti et al., 2023a). Family support is defined as the attitudes and behaviours of the family in supporting its members which can take the form of emotional support (e.g., care and comfort), informational support (e.g., guidance and advice), or instrumental support (e.g., practical and tangible assistance).(Chan et al., 2023) Family members are expected to be the most important caregivers for people experiencing schizophrenia. Schizophrenia has a significant negative impact on patients and their families. Family caregivers of people with schizophrenia experience high levels of stress and also have high levels of burden. This high burden includes physical discomfort, disrupted habits or routines tension, violence, chronic grief, enormous stigma, role changes, social withdrawal, and financial/career difficulties, usually with a lack of support resources.. (Chen et al., 2019) This article will discuss the philosophical study of family support in caring for schizophrenic patients based on ontology, epistemology and axiology.

### 1. Ontological study of family support in caring for schizophrenia patients

The concept of Ontology in the context of family support for schizophrenia patients can be understood as the important role of the family in maintaining the existence and identity of patients, as well as maintaining their mental and emotional stability. The family’s presence in the patient’s care is not only important for their social existence, but also in helping patients rebuild a more consistent and coherent perception of their world. In this regard, the family ontology becomes vital in shaping a more accessible reality structure for patients, helping them to overcome the challenges of schizophrenia. Family support is indispensable in the care of schizophrenia patients. This support includes medication supervision, ongoing care, and empowerment. However, caregivers face great emotional, financial, and social burdens. Support programmes and family-centred treatment approaches can reduce these challenges, leading to better outcomes for patients and caregivers. Families are critical in overseeing treatment adherence and are also crucial to the patient’s recovery process. (Purba et al., 2020) Providing ongoing care and optimising support systems is essential. Families assist in daily activities and ensure the patient’s needs are met. In addition, family support can also include strengthening the patient by involving them in social activities and helping develop their social skills. (Fitryasari et al., 2024b). The challenges faced by family caregivers are that caregivers often experience significant emotional and psychological distress due to the demanding nature of caregiving. Financial pressures and limited social interaction add to the burden, making it difficult for carers to manage their responsibilities effectively. (Lekganyane, 2020) There is often inadequate support from the healthcare system and other family members, which exacerbates the challenges of caregivers (Asgari et al., 2023). Family support given to caregivers as well as to patients provides better outcomes for patients. Effective family support leads to better outcomes for patients, including increased independence and improved daily functioning.( Rohmi et al., 2020). Adequate social support significantly reduces the burden on caregivers, improving their ability to provide care (Sustrami, Yusuf, Fitryasari, Efendi, et al., 2023)

### 2. Epistemiological study of family support in caring for schizophrenia patients

Schizophrenia is a chronic disorder that often requires long-term treatment. Family support is essential to ensure that patients have the emotional and practical resources necessary to navigate daily life. Without such support, patients may feel lonely, isolated, and demoralised to undergo treatment, which can worsen their symptoms. Family provides continuity and emotional stability that often cannot be found in a purely professional medical relationship. A patient with schizophrenia can be severely affected by changes in medical care, but family can offer more constant stability, allowing the patient to feel more secure and accepted. Low knowledge and high family burden in caring for the patient were the most common factors experienced by families of patients with schizophrenia. This shows that these two factors can be the cause of relapse in schizophrenia patients so it needs to be considered by nurses.(Suryani et al., 2019) Family support plays an important role in the treatment and recovery of individuals with schizophrenia as families are the primary carers for individuals with schizophrenia, providing essential practical and emotional support. There is an association between family emotional expression, infrastructure and health workers with the family’s ability to care for patients with schizophrenia, although these three variables are not predictors.(Iswanti et al., 2023b) Caregivers experience significant physical, psychological, and economic burdens, including stress, anxiety, and reduced quality of life (Chen et al., 2019). Family support is essential for patient engagement in treatment and recovery, especially in the early stages of psychosis (Purba et al., 2020)(Demarais et al., 2024). Effective family support is associated with improved quality of life for patients and caregivers (Setiawati et al., 2021). Families also often experience challenges in providing care support to patients with schizophrenia. Stigmatisation of mental illness can lead to social isolation and additional psychological distress for caregivers.(Fatima & Tariq, 2022). Family and social support also mediate the intensity of healing indicators and the decline in disease indicators, and the source of support is critical to the course of the disease. (Izydorczyk et al., 2019) Caring for someone with schizophrenia is a very challenging task, and can leave family members feeling exhausted or even depressed. There are several barriers perceived by families in caring for these schizophrenic patients, namely inefficient support resources including the subcategories of inadequate support by family members, inadequate support by the healthcare system and financial difficulties. Other barriers include limited public knowledge about mental disorders including social stigmatisation and social rejection. The destructive nature of schizophrenia is also a barrier including gradual loss of abilities and gradual patient passivity.(Asgari et al., 2023). Family members should endeavour to learn more about schizophrenia - its symptoms, ways of managing it, and available therapies and medications. Better knowledge helps the family to be more thoughtful in providing support and also to avoid misunderstandings or missteps in responding to the patient’s behaviour. Structured family interventions, such as psychoeducation and support programmes, have been shown to improve patient and caregiver outcomes. (Bademli & Duman, 2011). Family support is indispensable in the care of individuals with schizophrenia, yet caregivers face many challenges that can affect their well-being and effectiveness. Addressing these challenges through targeted interventions and better healthcare policies can significantly improve support systems for patients and their families.

### 3. Axiological study of family support in caring for schizophrenia patients

Family support in caring for schizophrenia patients is very important as it contains a number of fundamental values that lead to individual well-being and meaningful social relationships. From an axiological perspective, family support is a manifestation of the values of compassion, human dignity, social solidarity, responsibility, meaningful life, and justice. All these values work together to ensure that patients not only get physical care, but also feel valued, accepted, and empowered in their lives. Family support enables patients to find meaning in their lives, maintain social integrity, and provide a much-needed sense of security in their healing process. In this context, the family is not only a provider of support, but also a custodian of human values that help patients live a life full of dignity despite the challenges. Key Values and Benefits of Family Support Providing emotional support is important for the psychological well-being of patients with schizophrenia. This support helps reduce psychological stress and emotional burden on the patient, fostering a more stable environment for recovery.(Purba et al., 2020) Family support helps reduce the social stigma associated with schizophrenia by providing a supportive and understanding environment, families can improve patients’ social integration, which is crucial for their overall quality of life.(Asgari et al., 2023). Practical support, such as assistance with daily activities and medication management, is essential. Studies show that family instrumental support significantly improves medication adherence, which is crucial for effectively managing schizophrenia.( Rohmi et al., 2020) Financial support from family can ease the economic burden associated with long-term care for schizophrenia patients. This support is often needed to cover the cost of medication and other related expenses, which can be substantial.(Chen et al., 2019) Family support is invaluable in the care of patients with schizophrenia, providing emotional, practical, and financial benefits that significantly impact patient recovery and caregiver well-being. Addressing the challenges faced by caregivers through structured support programmes and policies is critical to improving the overall quality of care for individuals with schizophrenia.

## Conclusion

Based on a review of 10 articles discussing family support in caring for schizophrenia patients, it was found that family support is very important in the care of schizophrenia patients. There are several barriers that prevent families from accessing necessary resources and support, including stigma, lack of information, and limited contact with the healthcare team. Family support provided to caregivers as well as to patients resulted in better outcomes for patients including increased independence and improved daily functioning, reduced caregiver burden and improved caregiver ability to provide care.

## Data Availability

All data produced in the present work are contained in the manuscript

